# Predicting the risk of diabetes complications using machine learning and social administrative data in a country with ethnic inequities in health: Aotearoa New Zealand

**DOI:** 10.1101/2023.08.18.23294287

**Authors:** Nhung Nghiem, Nick Wilson, Jeremy Krebs, Truyen Tran

## Abstract

**Background:** In the age of big data, linked social and administrative health data in combination with machine learning (ML) is being increasingly used to improve prediction in cardiovascular diseases (CVD). We aimed to apply ML methods on extensive national-level health and social administrative datasets to predict future diabetes complications by ethnicity.

**Methods:** Five ML models were used to predict CVD events among all people with known diabetes in the population of New Zealand, utilizing national-level administrative data at the individual level.

**Results:** The Xgboost ML model had the best predictive power for predicting CVD events three years into the future among the population with diabetes. The optimization procedure also found limited improvement in AUC by ethnicity. The results indicated no trade-off between model predictive performance and equity gap of prediction by ethnicity. The list of variables of importance was different among different models/ethnic groups, for examples: age, deprivation, having had a hospitalization event, and the number of years living with diabetes.

**Discussion and conclusions:** We provide further evidence that ML with administrative health data can be used for meaningful future prediction of health outcomes. As such it could be utilized to inform health planning and healthcare resource allocation for diabetes management and the prevention of CVD events. Our results may suggest limited scope for developing prediction models by ethnic group and that the major ways to reduce inequitable health outcomes is probably via improved delivery of prevention and management to those groups with diabetes at highest need.

## Background

People living with diabetes have a higher risk for cardiovascular disease (CVD) events than the general population.^(1)^ According to the Global Burden of Disease Study 2017, CVD is the leading cause of death in the world.^(2)^ Some treatments for CVD can be very expensive and cumulatively account for a large proportion of total health system costs.^(3, 4)^ Therefore predicting CVD events among people with diabetes is desirable for health system planning. In addition, diabetes and CVD events are more prevalent in some ethnic groups than the others,^(5, 6)^ and this needs to be taken into account in health outcome prediction. In Aotearoa New Zealand (NZ), diabetes and CVD are the leading causes of premature death and disease burden, and are major sources of health inequities for Māori, Pasifika, and Asian populations due to socio-economic, cultural and health system factors.

There is strong evidence around the prevention of diabetes complications, such as controlling glucose levels, hypertension, dyslipidemia and smoking cessation.^(7)^ However, there are factors at a system-level, which compromise the ability to act upon this evidence and care for populations, such as socio-economic status, medication costs, and access to healthcare.^(8)^ Health inequities in NZ have long been recognized, yet little improvement has been achieved over the last 20 years or more.^(9)^ More urgent action and policy interventions beyond the health system are needed to reduce health burdens in marginalized populations.^(9)^

The NZ Government, similar to the governments in Scandinavian countries, United Kingdom, and Australia, holds a large amount of data from patient interactions with the healthcare system.^(10)^ This is in addition to extensive other individual data such as census, immigration, and justice data, which can be linked at an individual level. These data are high-dimensional, very extensive and impossible to explore by clinicians or health systems decision makers manually.

Machine learning (ML) method has emerged as a promising new technique to model disease risk prediction in an era of large datasets.^(11–14)^ It consists of a large number of alternative methods including classification trees, random forest, neural networks, support vector machines, and lasso and ridge regression. For studies where the primary goal is to predict the occurrence of an outcome, this technique produces a more flexible relationship among the predictor variables and the outcome.^(11)^ ML can accommodate non-linear relationships while overcoming the over-fitting issues in the traditional regression models.^(15–17)^ In fact, the emerging evidence suggests that ML significantly improves accuracy of CVD risk prediction compared to the traditional regression models.^(8, 15, 17, 18)^

There are a large number of prediction models that have been developed for CVD events among people with diabetes in the clinical setting,^(8)^ including both traditional regression and ML methods.^(1, 19)^ These models generally utilize rich clinical information or features (eg, body mass index, smoking status, biomarkers) extracted from electronic medical records or clinical trials. However, while these models are important for risk prediction at a clinical level, they are not easily deployed at the population level in order to reduce systemic barriers to improve diabetes management. In contrast, linked social and administrative health data consists of records collected on diagnoses, medications, and demographics generated through the provision of health services by governments. These data have become increasingly available to assess population health,^(10, 20)^ and they represent a valuable resource for automated analytic approaches to improve the efficiency and effectiveness of primary and secondary health prevention efforts.^(8)^

Given this background, the overall aim of this research was to: 1) use ML models to predict CVD events over a three-year period for the NZ adult population with diabetes using a broad range of routinely collected health and social administrative data; and 2) assess the performance of ML models on different ethnic groups in NZ to determine the relevance to reducing health inequities.

## Methods

### Datasets

We used linked health and social administrative data from the Stats NZ (SNZ) Integrated Data Infrastructure (IDI). This is a research database that links a broad and diverse collection of administrative and survey datasets from health, income, benefits and social services, education, justice, housing, and communities. All individual data across different datasets were linked through a unique identifier code.

The first dataset was the Census 2013 to identify individuals’ smoking status, language spoken, employment status and other demographic information. The second dataset, the diabetes complications dataset from the Ministry of Health (MoH chronic condition table), contains information about healthcare users in the population cohort who have been diagnosed with one or more of eight chronic conditions (eg, coronary heart disease, stroke, diabetes, cancer, and gout). We used this dataset to identify people with CVD and diabetes, and other chronic diseases in 2013.^(21)^ In order to identify individuals on CVD preventive pharmacotherapy, we used pharmaceutical data from 2013, but with no history of a CVD event (ie, individuals that had: (i) none of the conditions in the MoH chronic conditions table; or (ii) did have one of these conditions, but who had no prior identified CVD condition). This dataset contains claim and payment information from pharmacists for subsidized dispensings. Finally, we used the IDI Population Explorer dataset (2013), which has indicators for receipt of social security benefit, use of social housing, and major life events (ie, getting divorced/separated when this was officially documented) in 2013.^(4)^ Patients or the public were not involved in the design, or conduct, or reporting, or dissemination plans of our research.

### Study population

The whole population of NZ who were in the residential population in 2013, and who had been diagnosed with diabetes in Virtual Diabetes Registry in the period of January 2001 to December 2013 ^(22)^ but with no prior CVD, were followed throughout to 2018. In order to identify people with diabetes, any CVD complications they had, and their social characteristics, we used the International Classification of Diseases (ICD) for identifying diabetes and CVD complication events.^(3, 23)^ The definition for CVD was based on the following ICD-10 codes: stroke (I60-I64; G45-G46), and coronary heart disease (ICD-10-AM: I20-I25).^(22)^

Only people who were in both the Census 2013 and the IDI estimated resident population in 2013, and had diabetes but did not have diagnosed CVD, were included in the analysis. We also further restricted the population to people aged between 30 to 74 years old as per other NZ work in CVD risk prediction.^(20)^ All observations with missing age and sex information were excluded from the analysis but these were very infrequent. Steps to extract the study population were presented in Figure 1.

**Figure 1.**
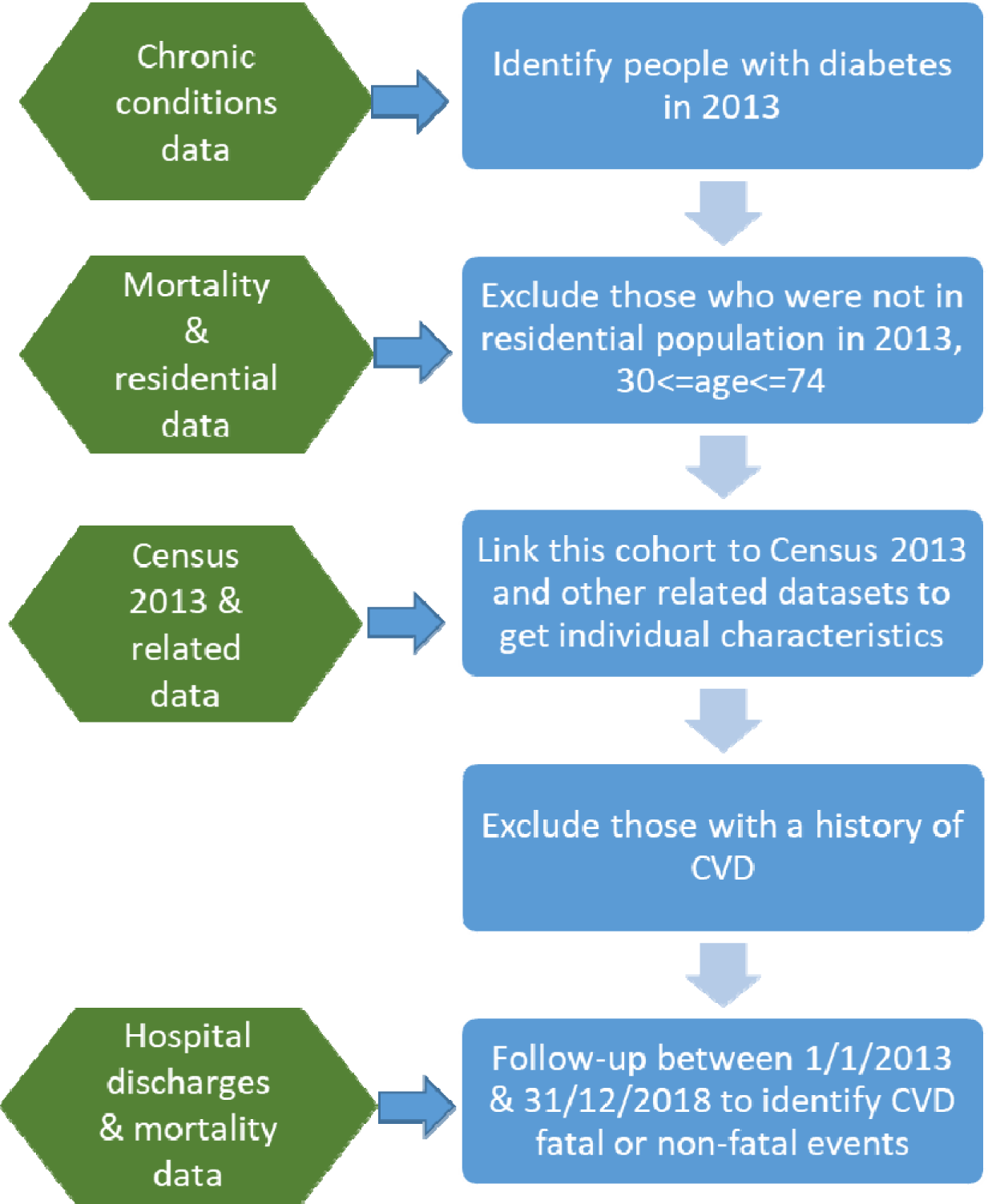
Steps to extract the study population from the linked administrative and health data.

### Outcome

This included the risks of developing CVD over a three-year period. The dependent variable was a binary outcome whether a CVD event (either fatal or non-fatal) had or had not occurred for an individual with diabetes during three-year periods between 1/1/2013-31/12/2015; and 1/1/2016-31/12/2018.

### Variables

The linked health and administrative datasets allow us to examine the individual-level impact of not only the health indicators (eg, diabetes, smoking status); but also other demographic characteristics (age, sex, ethnicity (self-identified), immigration status); social background variables (eg, housing conditions, social security benefits and language spoken); and potential stress indicators (via employment).

In addition, the following conditions were added to the predictor variables: any hospital event between 2001 and 2013 for dementia, asthma, chronic kidney disease, and total hospital events for any condition. Disease ICD-10 codes were extracted from the MoH Burden of Disease Report 2016.^(24)^ These conditions were added to the predictor variable list based on the literature for conditions associated with CVD.^(25–27)^

### Data pre-processing

The data were randomly divided into 80% training and 20% test. Each individual could only be in either training or test data.

### Data subsets by time period and by ethnicity

We split out datasets into a study dataset with a three-year follow-up from 2013-2015 and a validation dataset from 2016-2018.^(8)^ We also created datasets by ethnicity, in particular: the whole NZ population with diabetes, Asian population with diabetes, Māori population with diabetes and Pasifika population with diabetes.

### Model development and evaluation

We used ML models, such as logistic regression, decision trees, random forest, neural network, and Xgboost to predict CVD complications.^(28, 29)^ Following Zafar et al,^(30)^ two fold-cross validations were performed on the training data. Parameter tuning was performed using area under the receiver operating curve (AUC) as an evaluation matrix. Models were coded and analyzed in R version 3.3.0. All ML models were trained using the same training datasets and tested on the same test datasets to allow comparison of their predictive power. The main indicator AUC was used to evaluate the predictive performance of the ML models.

### Model optimization

ML models were trained to maximize the AUC indicators, either for the whole NZ population with diabetes or for a particular ethnic group (eg, Asian) as per Figure 2. The aim was to improve model performance for a particular ethnic group in order to understand fairness in disease prediction. Our measure is somewhat similar to the group fit measurement employed by McGuire et al.^(31)^ However, while these authors used group fit for the total payment ratio received for groups by health condition (cancer, heart health, diabetes and mental health), our group fit was AUC by ethnicity. Furthermore, McGuire et al. set up a constraint on the group fit measurement (ie, the total payment ratio equals one reflecting a balance between budgeted and actual health expenditures), but we optimized our group fit level by ethnicity through parameter tuning.

**Figure 2.**
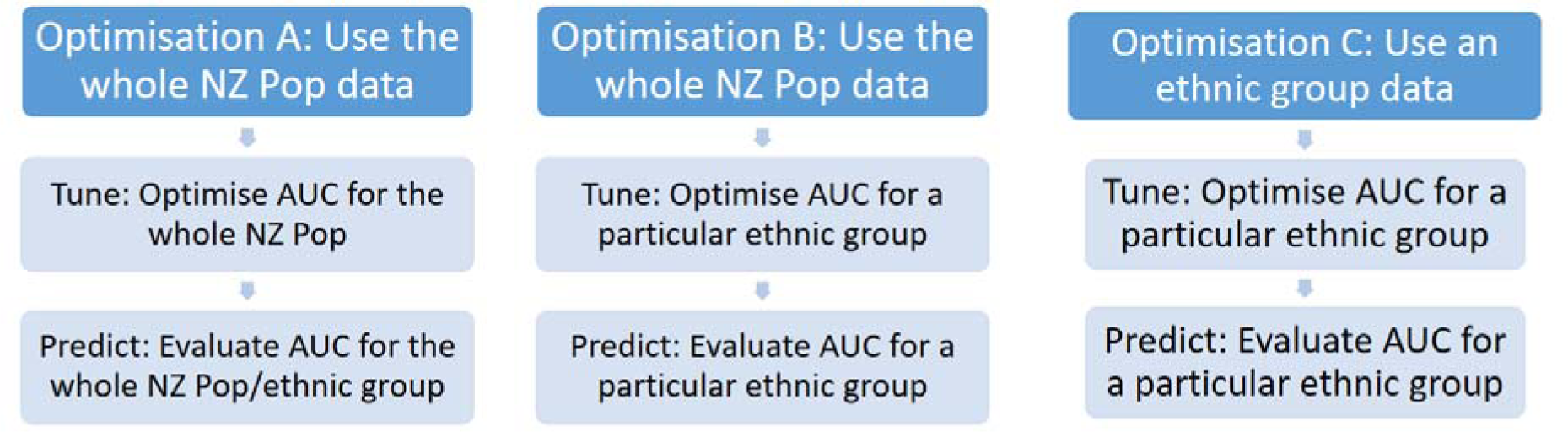
Optimization scenarios with different data subsets and evaluation indicators for populations with diabetes.

## Results

### Descriptive results

As shown in Table 1, there were approximately 146,000 NZ residents with diabetes who were aged 30-74 and with complete data on basic demographic information: age and sex. There were less than 0.5% of observations having missing ethnicity data, and less than 10% missing smoking status data. All observations with missing data other than age and sex were included in the analyses and were implicitly treated as missing data. Table 2 presents CVD incidence rates among people with diabetes by various predictors, in particular age, sex, ethnicity, deprivation decile, smoking status, and employment status.

**Table 1:**
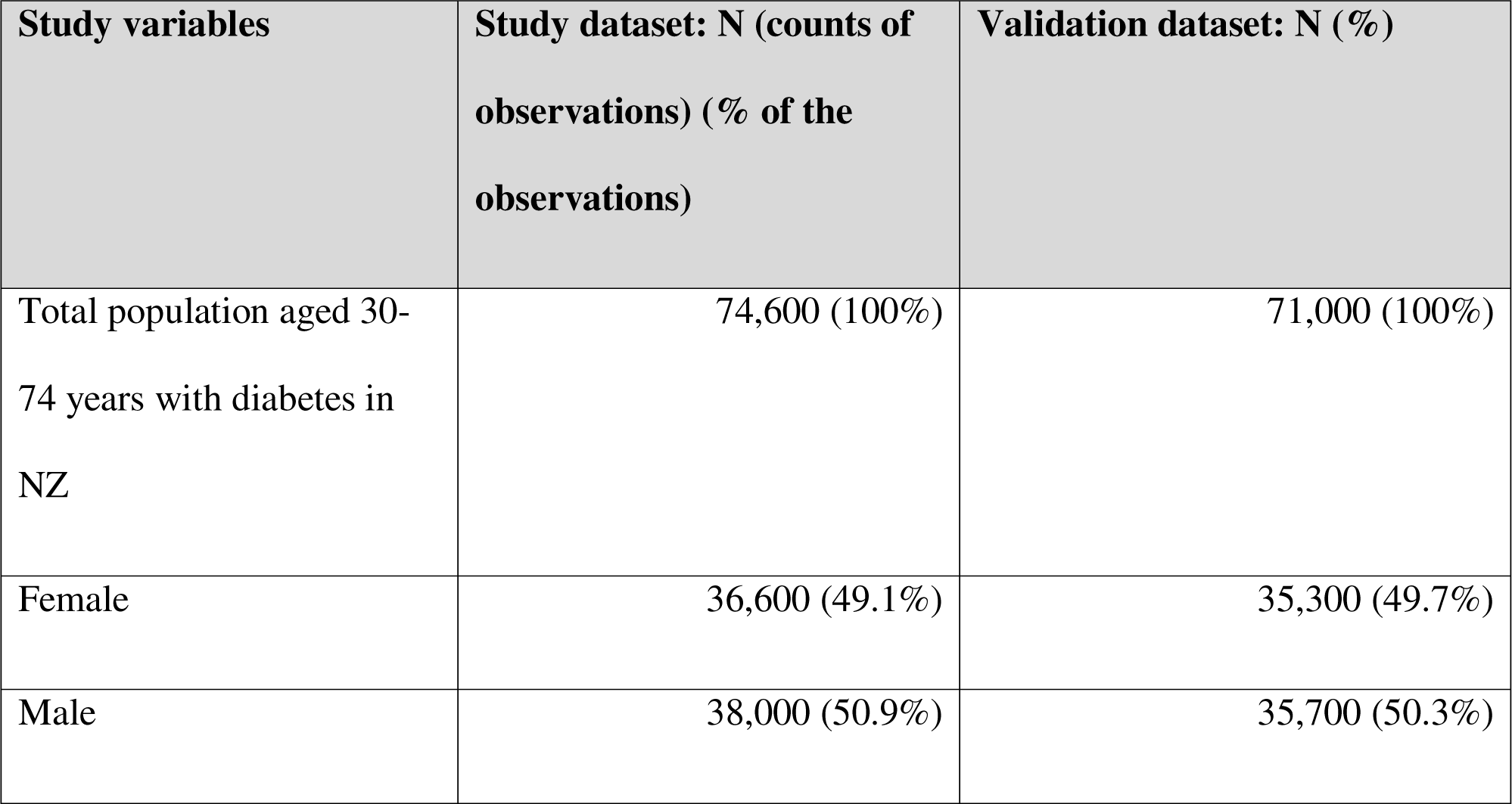
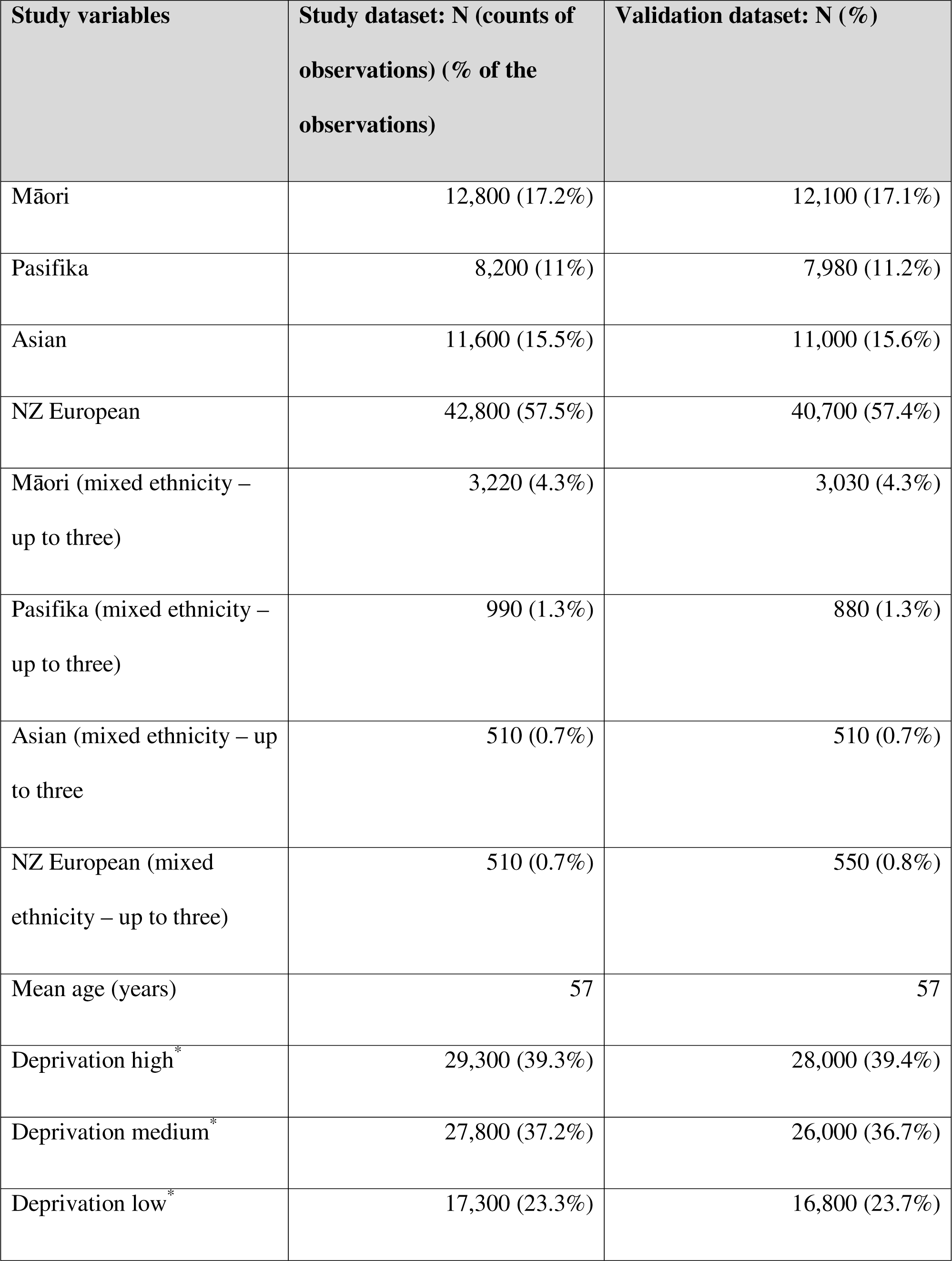
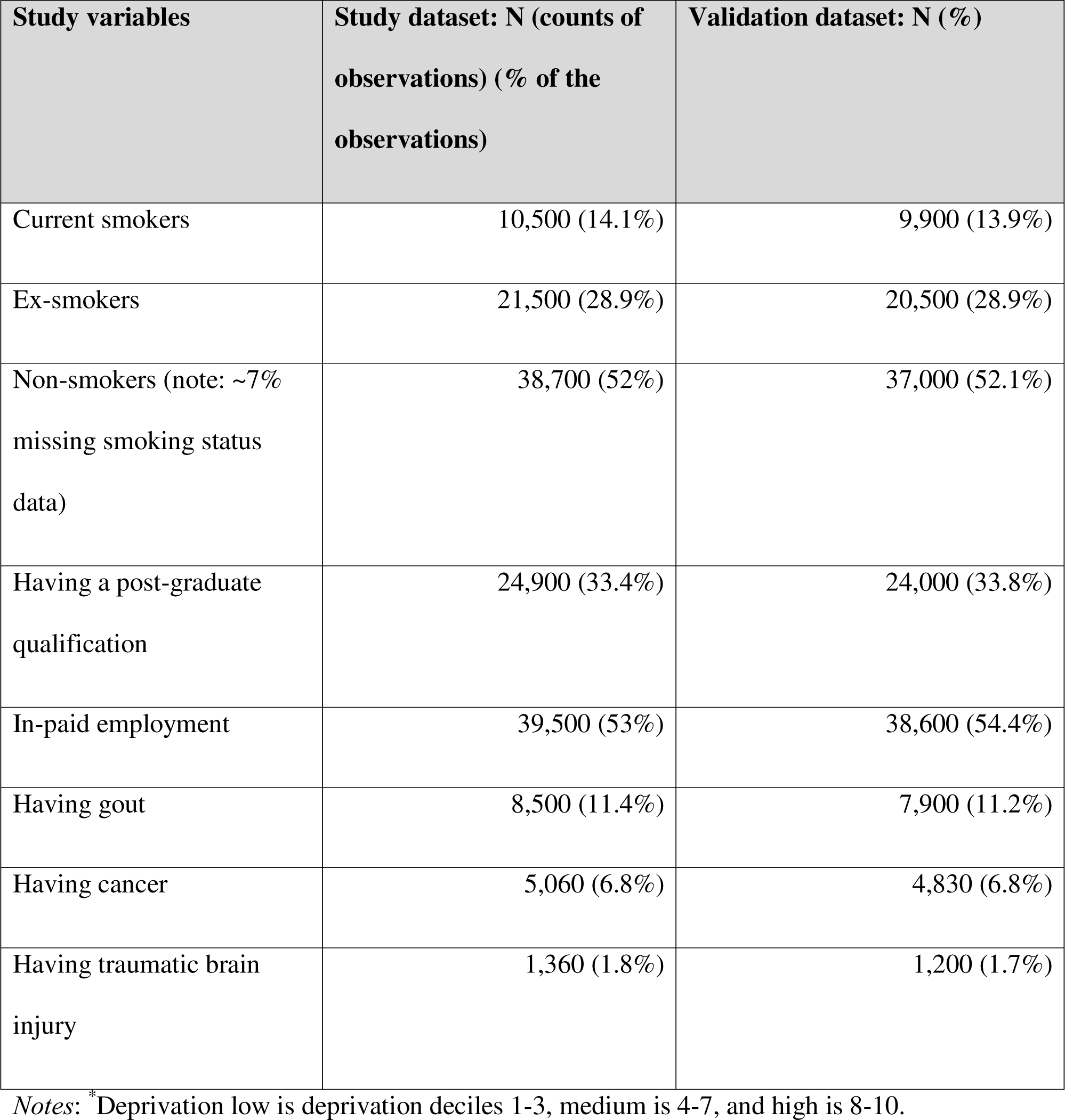
Descriptions of variables included in the analysis in both study and validation datasets.

| Study variables | Study dataset: N (counts of observations) (% of the observations) | Validation dataset: N (%) |
| --- | --- | --- |
| Total population aged 30-74 years with diabetes in NZ | 74,600 (100%) | 71,000 (100%) |
| Female | 36,600 (49.1%) | 35,300 (49.7%) |
| Male | 38,000 (50.9%) | 35,700 (50.3%) |

| **Study variables** | **Study dataset: N (counts of observations) (% of the observations)** | **Validation dataset: N (%)** |
| --- | --- | --- |
| Māori | 12,800 (17.2%) | 12,100 (17.1%) |
| Pasifika | 8,200 (11%) | 7,980 (11.2%) |
| Asian | 11,600 (15.5%) | 11,000 (15.6%) |
| NZ European | 42,800 (57.5%) | 40,700 (57.4%) |
| Māori (mixed ethnicity – up to three) | 3,220 (4.3%) | 3,030 (4.3%) |
| Pasifika (mixed ethnicity – up to three) | 990 (1.3%) | 880 (1.3%) |
| Asian (mixed ethnicity – up to three) | 510 (0.7%) | 510 (0.7%) |
| NZ European (mixed ethnicity – up to three) | 510 (0.7%) | 550 (0.8%) |
| Mean age (years) | 57 | 57 |
| Deprivation high<sup>*</sup> | 29,300 (39.3%) | 28,000 (39.4%) |
| Deprivation medium<sup>*</sup> | 27,800 (37.2%) | 26,000 (36.7%) |
| Deprivation low<sup>*</sup> | 17,300 (23.3%) | 16,800 (23.7%) |
| Current smokers | 10,500 (14.1%) | 9,900 (13.9%) |
| Ex-smokers | 21,500 (28.9%) | 20,500 (28.9%) |
| Non-smokers (note: ~7% missing smoking status data) | 38,700 (52%) | 37,000 (52.1%) |
| Having a post-graduate qualification | 24,900 (33.4%) | 24,000 (33.8%) |
| In-paid employment | 39,500 (53%) | 38,600 (54.4%) |
| Having gout | 8,500 (11.4%) | 7,900 (11.2%) |
| Having cancer | 5,060 (6.8%) | 4,830 (6.8%) |
| Having traumatic brain injury | 1,360 (1.8%) | 1,200 (1.7%) |

Notes: \* Deprivation low is deprivation deciles 1-3, medium is 4-7, and high is 8-10.

**Table 2.**
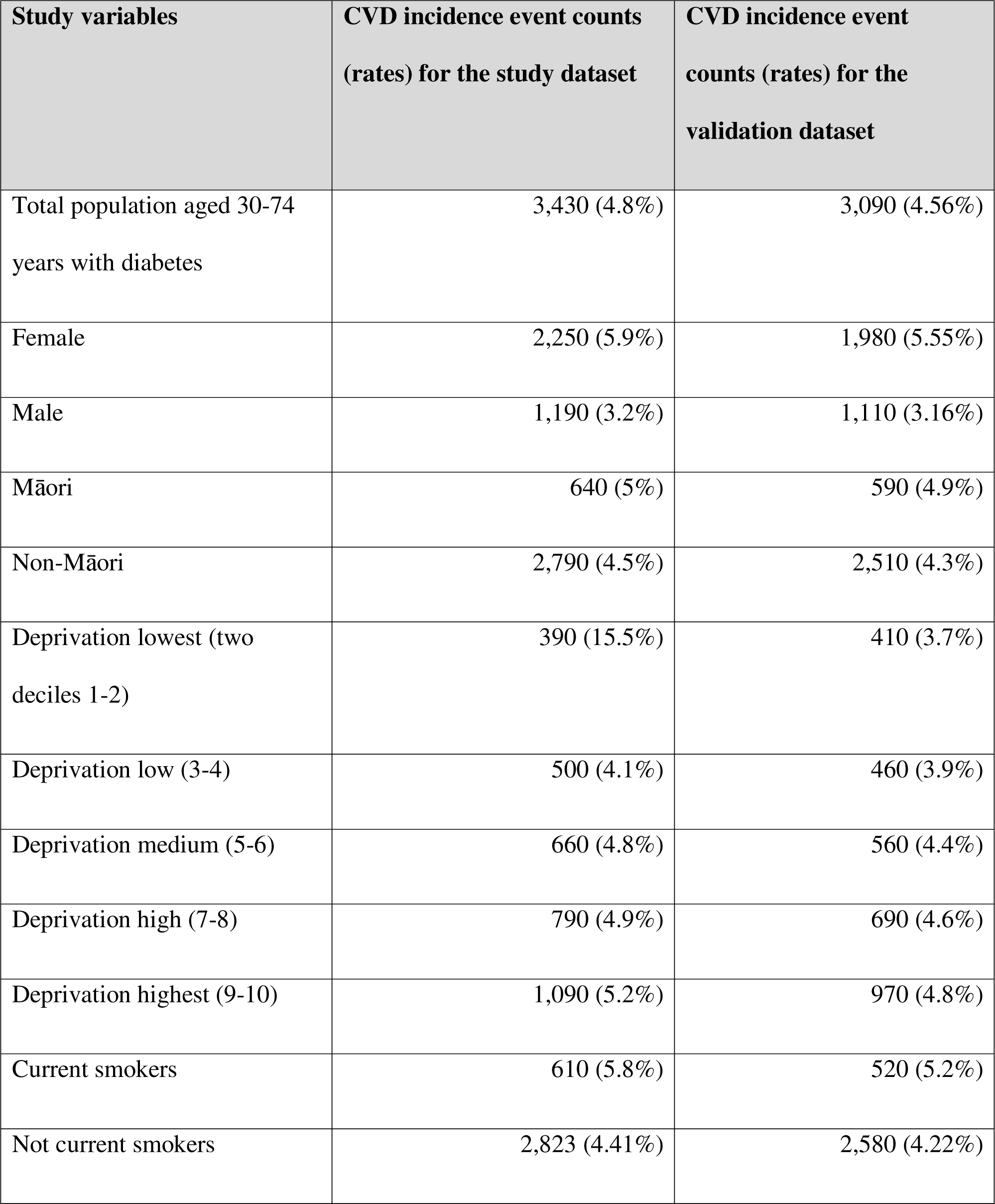
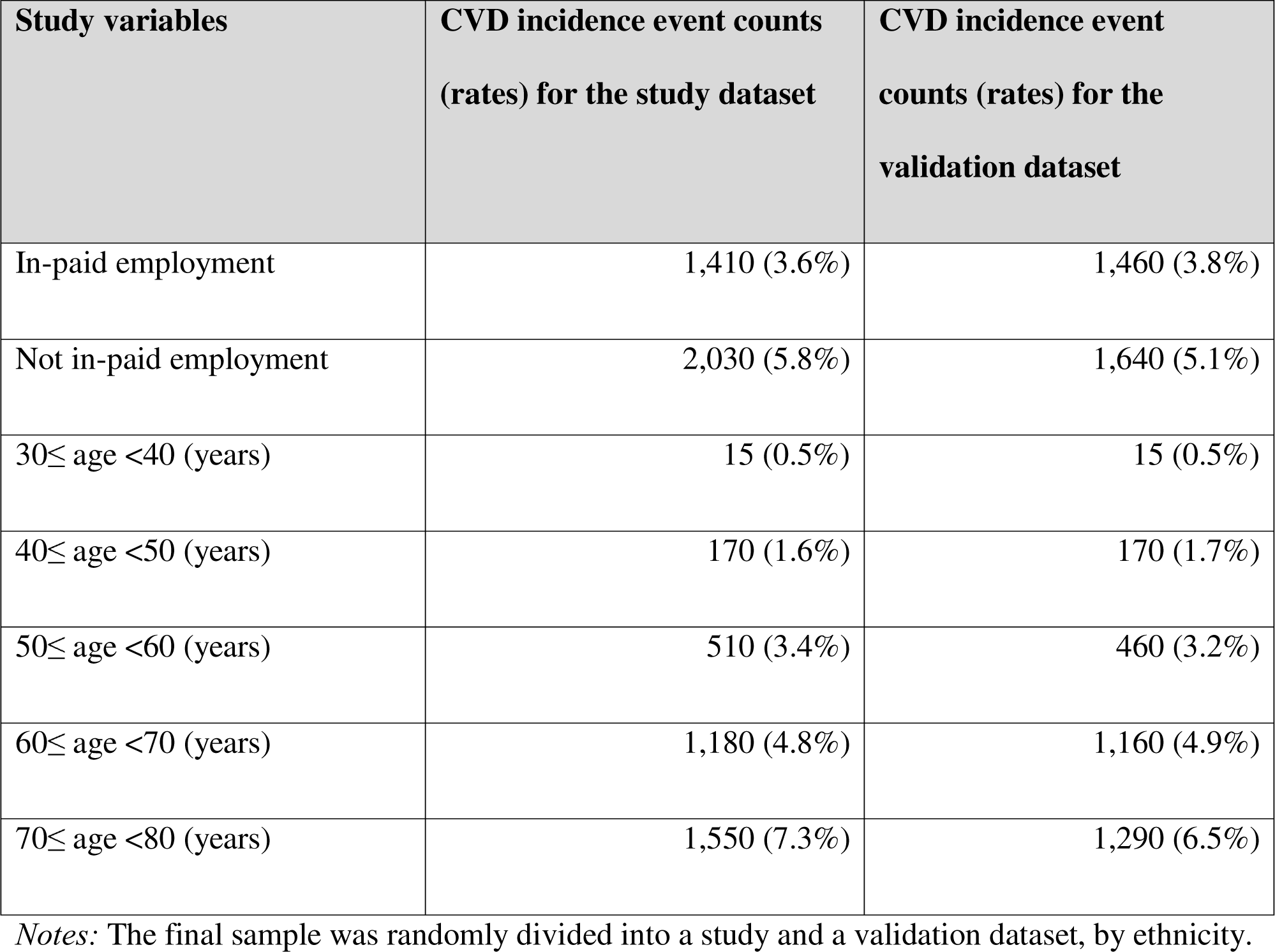
CVD incidence counts and rates during the three-year follow-up period by demographic information in the study population.

| **Study variables** | **CVD incidence event counts**<br>**(rates) for the study dataset** | **CVD incidence event**<br>**counts (rates) for the**<br>**validation dataset** |
| --- | --- | --- |
| Total population aged 30-74<br>years with diabetes | 3,430 (4.8%) | 3,090 (4.56%) |
| Female | 2,250 (5.9%) | 1,980 (5.55%) |
| Male | 1,190 (3.2%) | 1,110 (3.16%) |
| Māori | 640 (5%) | 590 (4.9%) |
| Non-Māori | 2,790 (4.5%) | 2,510 (4.3%) |
| Deprivation lowest (two<br>deciles 1-2) | 390 (15.5%) | 410 (3.7%) |
| Deprivation low (3-4) | 500 (4.1%) | 460 (3.9%) |
| Deprivation medium (5-6) | 660 (4.8%) | 560 (4.4%) |
| Deprivation high (7-8) | 790 (4.9%) | 690 (4.6%) |
| Deprivation highest (9-10) | 1,090 (5.2%) | 970 (4.8%) |
| Current smokers | 610 (5.8%) | 520 (5.2%) |
| Not current smokers | 2,823 (4.41%) | 2,580 (4.22%) |
| In-paid employment | 1,410 (3.6%) | 1,460 (3.8%) |
| Not in-paid employment | 2,030 (5.8%) | 1,640 (5.1%) |
| 30≤ age <40 (years) | 15 (0.5%) | 15 (0.5%) |
| 40≤ age <50 (years) | 170 (1.6%) | 170 (1.7%) |
| 50≤ age <60 (years) | 510 (3.4%) | 460 (3.2%) |
| 60≤ age <70 (years) | 1,180 (4.8%) | 1,160 (4.9%) |
| 70≤ age <80 (years) | 1,550 (7.3%) | 1,290 (6.5%) |

*Notes:* The final sample was randomly divided into a study and a validation dataset, by ethnicity.

Table 3 presents model performance by ethnicity across two time periods (2013-2015 and 2016-2018). Models were trained using data for the whole NZ population aged 30-74 years with diabetes in 2013-2015, were optimized for the indicator (AUC) for this population, and were predicted by ethnic group. When there was no change in time period (that is training and test datasets were in the same period), results suggested that Xgboost models outperformed all other ML models in term of preventing future CVD events – based on AUC, across ethnicity and time periods. In particular, the average AUC by time period was 0.74 for the whole NZ population with diabetes, and similarly for other populations: for 0.74 for the Asian (0.74), Māori (0.76), and Pasifika population (0.73). Compared to the random forest (RF) models, the prediction by Xgboost models was improved by 6.4% (0.74 vs 0.70) for the whole NZ population with diabetes and 10% (0.73 vs 0.66) for the Pasifika population.

**Table 3.** Model performance by ethnicity across two time periods (2013-2015, 2016-2018) (Models were trained using data for the whole NZ population with diabetes in 2013-2015, were optimized for the indicator (AUC) for this population, and were predicted by ethnic group)

| **Models\*** | **Model prediction (AUC) for the 2013-2015 period (current period)** | **Model prediction (AUC) for the 2016-2018 period (future period)** | **Average AUC for both time periods** | **Absolute AUC gaps between two time periods ($\leq 0$ means at least equal prediction)** |
| --- | --- | --- | --- | --- |
| Random forest (RF) all ethnic groups | 0.70 | 0.70 | **0.70** | **0.00** |
| RF Asian | 0.68 | 0.71 | 0.70 | -0.03 |
| RF Māori | 0.70 | 0.68 | 0.69 | 0.03 |
| RF Pasifika peoples | 0.66 | 0.68 | 0.67 | -0.02 |
| Xgboost all ethnic groups | 0.74 | 0.73 | **0.74** | **0.01** |
| Xgboost Asian | 0.74 | 0.76 | 0.75 | -0.01 |
| Xgboost Māori | 0.76 | 0.71 | 0.73 | 0.05 |
| Xgboost Pasifika peoples | 0.73 | 0.71 | 0.72 | 0.02 |

In term of predicting future CVD events, the RF models were quite similar to the Xgboost model for the whole NZ population with diabetes. But both models were slightly worse at predicting future events for Māori (absolute AUC gaps: 0.03 for RF and 0.05 for Xgboost, or about 4% and 7% worse, respectively). Both models seem to perform well for the Asian population in predicting future CVD events.

Model performance by ethnicity across subsets of data are presented in Table S3. Models were trained in 2013-2015, and were predicted by ethnic group in the same time period. There were three optimization scenarios as described in Figure 2. Results suggested that using all data (ie, all observations for the study population) for training and optimizing all data indicators (Optimization A) can improve the prediction compared to using sub-ethnicity data only (Optimization C) by 0.05 AUC (7.0%), 0.04 (5.2%), and 0.03 (4.8%) for Asian, Māori and Pasifika peoples, respectively, using the Xgboost model. Overall, Xgboost models benefited more from using population data than other models. With this current dataset, there were no benefits from optimizing ethnicity indicators (eg, building the optimal prediction so that it predicts best for Māori).

Table S4 shows gaps in model performance by ethnicity for the main indicator (AUC), using the study dataset in 2013-2015. The Xgboost models performed better in term of equity gaps, with an overall prediction improvement of 0.1% on average for sub-ethnic groups compared to the whole NZ population. The average improvement for the RF models was −2.3%, that is, the prediction for sub-ethnic groups was not as good as for the whole NZ population with diabetes. Table 4 presents a list of variables of importance generated by the RF models. Several main traditional risk factors for CVD were picked up (ie, being given higher ranking) by the RF models, in particular age, deprivation, and the number of years living with diabetes. Other socio-economic factors were also rated highly by the RF model, including: geographical area, income, deprivation, and occupation.

**Table 4.** Variables of Importance generated by the random forest model.

| **Rank** | **Variable of Importance** | **In a traditional regression model (ie, the NZ PREDICT equation)<sup>(1)</sup> (Yes/No)** |
| --- | --- | --- |
| **1** | Age | Yes |
| **2** | Geographical area (a smallest geographical area in NZ with code by regional council, territorial authority, ward, and area unit) | No |
| **3** | Having any hospitalization events | No |
| **4** | The number of years living with diabetes | Yes |
| **5** | Deprivation level | Yes |
| **6** | Having prescribed antiplatelet medicine | Yes |
| **7** | Income level | No |
| **8** | Having other chronic conditions prior to 2014, including cancer, gout, and traumatic brain injury. | No |
| **9** | Occupation (an occupation level that is not classified) | No |
| **10** | Having prescribed blood pressure lowering medicine | Yes |

## Discussion

### Interpretation of the main results

Our study demonstrated the feasibility of applying ML methods to administrative health data for public health planning, including taking into account fairness in terms of ethnicity. Our best model Xgboost can predict the three-year risk of CVD events in those with diabetes with an average AUC of 0.74. Our model was trained on test data for the NZ population with diabetes, which includes marked diversity by demographic and socio-economic variables. Our modelling was also validated in terms of the prediction of future events. The results suggested that the models generally performed slightly better for large groups of population. There seemed to be no trade-offs between the overall fit of the ML model and the fairness measurement in our analyses.

Our models performed reasonably well in comparison to the literature. In particular, the model to predict CVD risks among people with diabetes in the NZ setting from the 400,000-person primary care cohort study reported C-statistics of 0.73 and 0.69 for women and men, respectively.^(1)^ It should be highlighted that our models did not have rich clinical features (eg, BMI, SBP) as per the traditional risk prediction models, but was still able to produce comparable prediction results. We expected that if these clinical features were incorporated, the performance of the models would be improved. Our model’s performance was lower than the one developed in Canada^(8)^ (AUC of 0.74 vs 0.79) but this Canadian work had more data points. Of note is that, AUC and C-statistics are identical in the case of binary outcome, which is used in this study.^(32)^

Similar to the findings by the study in Canada, our variables of importance also picked up socio-economic factors as important variables in the prediction result.^(8)^ These variables include geographical area, income, occupation, and education level. The Canada study indicated that socio-demographic factors such as length of stay in Canada for immigrants and ethnic concentration in the area of residence, play an important role in model prediction.

### Study strengths and limitations

This study benefited from NZ having established some of the most comprehensive administrative health data holdings in the world, covering nearly the total population due to its universal healthcare system and digital government.^(10)^

Our ML models were validated against future time with no significant differences in model performance. These results were applied at both the total population level and the ethnic group level.

Nevertheless, this type of study is not currently easy to perform given that data used are held by the central government and the current computing infrastructure does not easily facilitate developing and running ML models on such large datasets. However, these constraints may ease with the expansion in size and speed of computing systems.

### Implications for health system

Administrative health data represent an enabler for automated analytic approaches to improve the efficiency and effectiveness of primary and secondary health prevention efforts, and address systemic barriers to diabetes care.^(8)^ Our findings suggest that ML can be capitalized to draw insights from administrative social and health data to improve health management and improve health equity.

While risk for CVD events among people with diabetes have been better managed in recent years, they remain a large burden because the incidence of diabetes continues to grow. Thus, there is a need to effectively prevent and manage diabetes complications at not only the individual patient level but also system levels. There was no trade-off between prediction performance and equity for other indicators; that is we can improve model prediction and reduce model performance gaps by ethnicity simultaneously. Furthermore, model training separately by ethnicity did not work well, so it appears best to use population data with ethnicity information, rather than train separate model for each ethnicity.

Even though our aim was to develop a prediction model for deployment at a population level, our variables of importance can still be further tested (ie, through a lasso logistic regression) to create a checklist to be used in the primary healthcare setting. Linked administrative and health databases typically have millions of records spread across multiple datasets making it highly challenging to work with. Moreover, predictive patterns inferred by the model at this scale can identify new trends or new risk factors at the population level. These variables may not be available in clinical prediction models as they generally exclude such types of features and mainly focus on health data for each patient. Thus the application of a ML model developed on administrative datasets to allocate resources and plan policies at a population level to improve diabetes complications outcomes could offer a data-driven approach to addressing health disparities.^(8)^

### Future research

With the improvement in computing power that allows processing a large amount of data, the number of features can be expanded to investigate yet unknown CVD risk factors in order to target public health or individual-level interventions. The methodology of this study could be applicable to other chronic diseases in NZ.

Future analysis may benefit from better accounting the possibility of misclassification in terms of ethnicity, such as the misclassification of Māori as a non-Māori,(33) in order to account for equity issues in NZ.

## Conclusions

We provide further evidence that ML with administrative health data can be used for meaningful future prediction of health outcomes. As such it could be utilized to inform health planning and healthcare resource allocation for diabetes management and the prevention of CVD events. Importantly, the ML model performance was only slightly different between ethnic groups in the NZ context and datasets. This may suggest limited scope for developing prediction models by ethnic group and that the major ways to reduce existing inequitable health outcomes is probably via improved delivery of prevention and management to those groups with diabetes at highest need.

## Declarations

### Ethics Approval and consent to participate

The study was approved by University of Otago ethics approval processes, reference number HD20/012. There were no participants directly involved in this study.

### Consent for publication

Not applicable.

### Availability of data and materials

Access to the anonymised data used in this study was provided by Stats NZ under the security and confidentiality provisions of the Statistics Act 1975. Only people authorised by the Statistics Act 1975 are allowed to see data about a particular person, household, business, or organisation, and the results in this paper have been confidentialised to protect these groups from identification and to keep their data safe.

### Authors’ contributions

NN, NW, JK, TT designed the study. NN, TT built the ML models. NN ran models and produced the results. NN, NW, JK, TT interpreted the results. NN wrote the first draft. NW, TT contributed significantly to the writing. All authors reviewed and approved the final draft of the manuscript.

## List of abbreviations

AUC: Area under the Receiver Operating Characteristics curve
CVD: Cardiovascular diseases
IDI: Integrated Data Infrastructure
ML: Machine learning
NZ: Aotearoa New Zealand
RF: Random Forest
SNZ: Stats New Zealand

## Acknowledgements

Access to the data presented was managed by Statistics NZ but under strict micro-data access protocols and in accordance with the security and confidentiality provisions of the Statistics Act 1975. Our findings are not Official Statistics. The opinions, findings, recommendations, and conclusions expresses are those of the authors, and not Statistics NZ and the University of Otago. The results are based in part on tax data supplied by Inland Revenue to Statistics NZ under the Tax Administration Act 1994. This tax data must be used only for statistical purposes, and no individual information may be published or disclosed in any other form, or provided to Inland Revenue for administrative or regulatory purposes. Any person who has had access to the unit record data has certified that they have been shown, have read, and have understood section 81 of the Tax Administration Act 1994, which relates to secrecy. Any discussion of data limitations or weaknesses is in the context of using the IDI for statistical purposes, and is not related to the data’s ability to support Inland Revenue’s core operational requirements.

We thank Sherri Rose for her useful advice on machine learning fairness at early stages of this research.

## APPENDIX

**Table S1.**
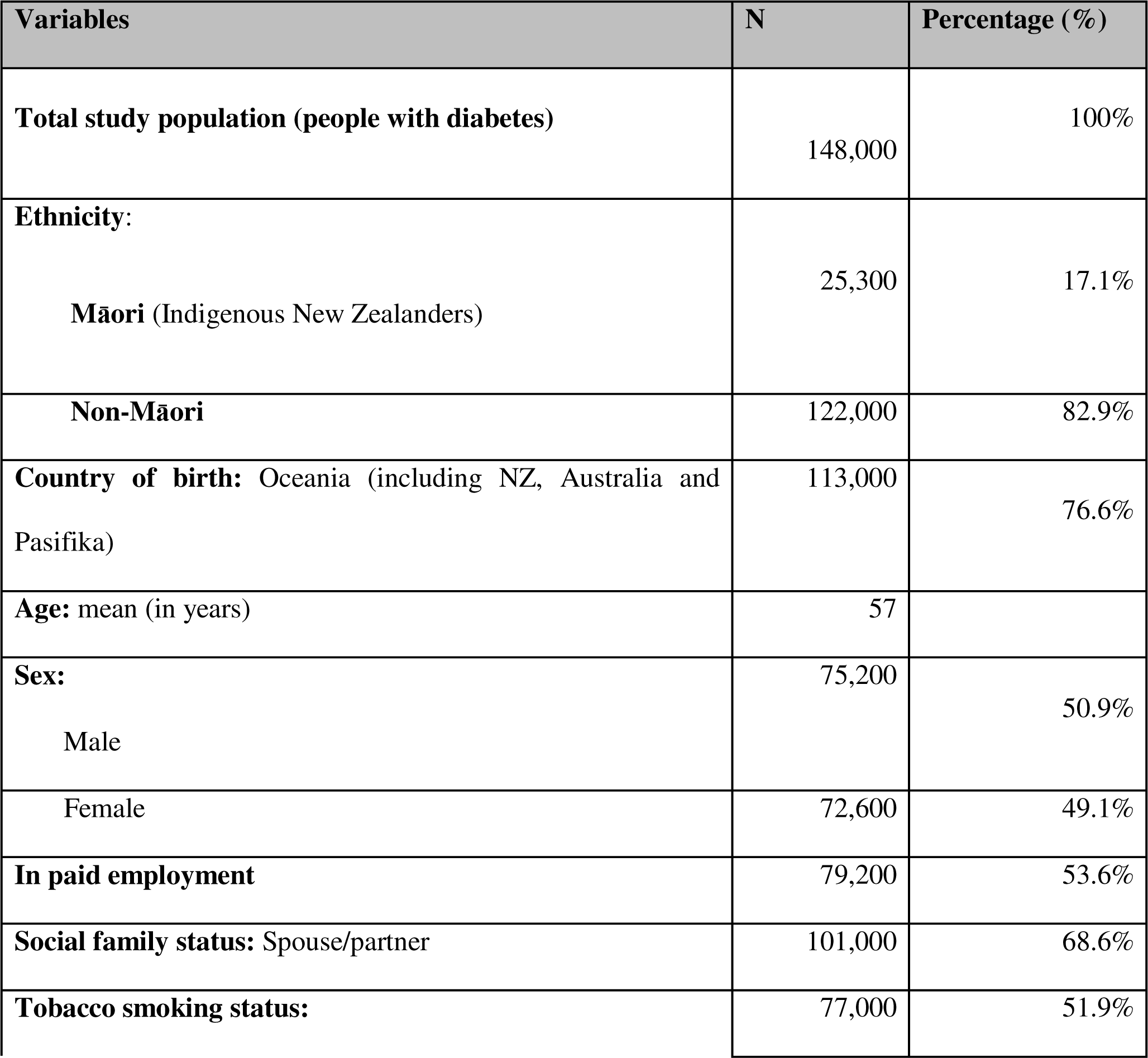
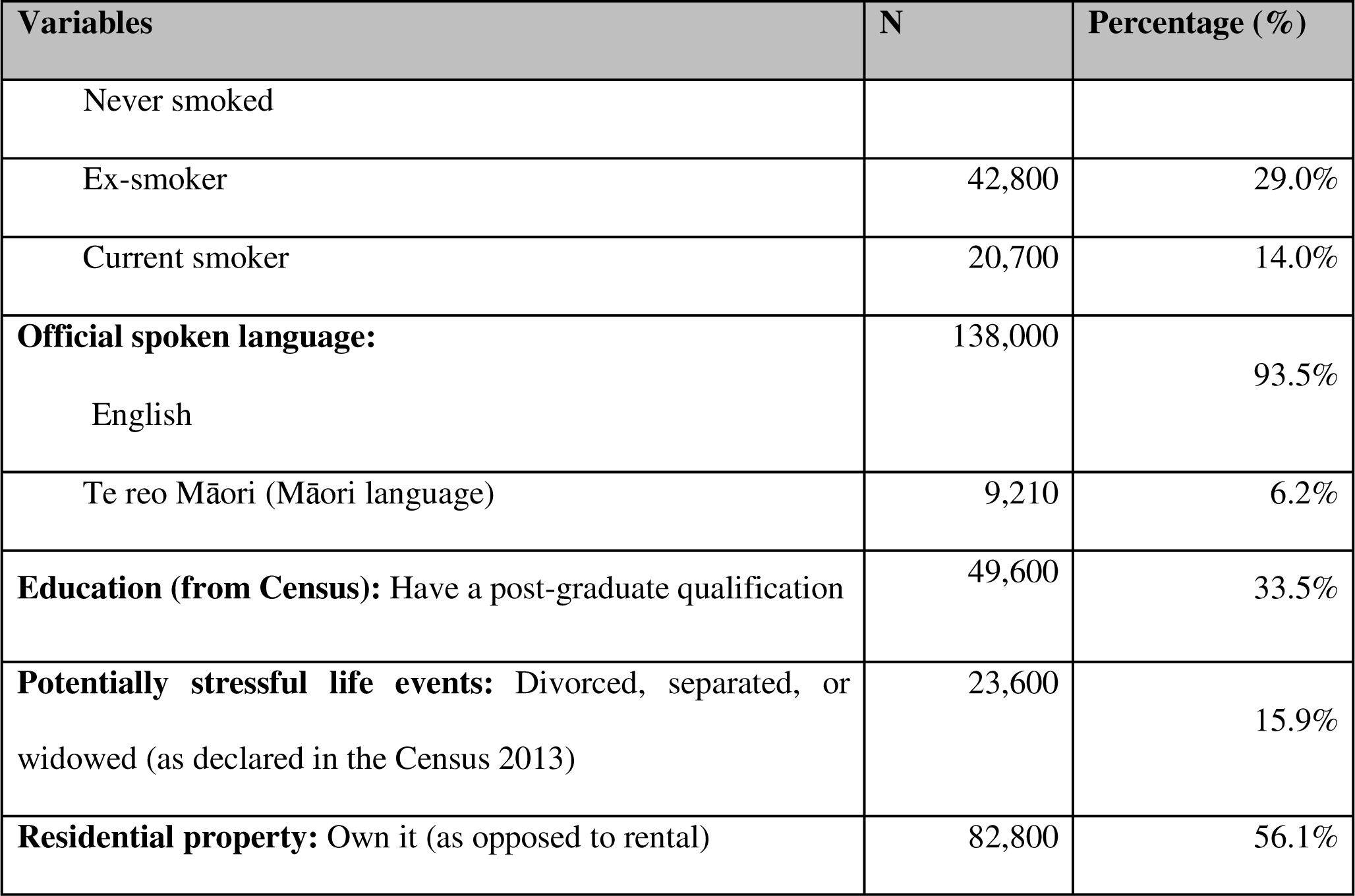
Description of socio-demographic and health variables used in the modeling. (The total population in 2013 before splitting between study and validation samples. The sub-categories might not add up to 100% due to rounding issues or missing data.)

**Table S2.** Gaps in model performance by ethnicity for other indicators.

| Model | Sensitivity | Specificity | F1 | Absolute gaps between ethnic group and the whole NZ population |  |  |
| --- | --- | --- | --- | --- | --- | --- |
|  |  |  |  | Sensitivity | Specificity | F1 |
| Random forest (RF) all ethnicity | 0.58 | 0.93 | 0.33 | N/A | N/A | N/A |
| RF Asian | 0.64 | 0.97 | 0.26 | 10% | 4% | -21% |
| RF Māori | 0.53 | 0.91 | 0.35 | -8% | -2% | 7% |
| RF Pasifika peoples | 0.37 | 0.93 | 0.17 | -35% | 0% | -50% |
| **Average RF** |  |  |  | **-11%** | **0%** | **-21%** |
| Xgboost all ethnicity | 0.29 | 0.77 | 0.39 | N/A | N/A | N/A |
| Xgboost Asian | 0.18 | 0.77 | 0.28 | -38% | 0% | -29% |
| Xgboost Māori | 0.29 | 0.77 | 0.39 | 2% | 0% | -1% |
| Xgboost Pasifika peoples | 0.23 | 0.77 | 0.34 | -21% | 0% | -14% |
| **Average Xgboost** |  |  |  | **-19%** | **0%** | **-15%** |

**Table S3.** Model performance by ethnic group across subsets of data. (Models were trained in 2013-2015 on training data, and were used for prediction by ethnic group in the same time period but on separate data)

| **Models** | **Average AUC across subsets of data** | **Optimization A: Models were trained using the whole NZ population data, and were optimized for the whole NZ population indicator (AUC)** | **Optimization B: Models were trained using the whole NZ population data, and were optimized by ethnic group's indicator** | **Optimization C: Models were trained using data by ethnic group, and were optimized by ethnic group's indicator** | **Absolute gaps in AUC between all data vs ethnic subset data** |
| --- | --- | --- | --- | --- | --- |
| Random forest (RF) all ethnic groups | NA | 0.70 | NA | NA | NA |
| RF Asian | 0.68 | 0.68 | 0.69 | 0.66 | 0.03 |
| RF Māori | 0.70 | 0.70 | 0.70 | 0.70 | 0.00 |
| RF Pasifika peoples | 0.65 | 0.66 | 0.66 | 0.63 | 0.03 |
| Xgboost all ethnic groups | NA | 0.74 | NA | NA | NA |
| Xgboost Asian | 0.72 | 0.74 | 0.74 | 0.69 | 0.05 |
| Xgboost Māori | 0.75 | 0.76 | 0.76 | 0.72 | 0.04 |
| Xgboost Pasifika peoples | 0.71 | 0.73 | 0.71 | 0.70 | 0.03 |
| **Average AUC** |  |  |  |  |  |
| RF |  | 0.69 | 0.68 | 0.66 | 0.02 |
| Xgboost |  | 0.74 | 0.74 | 0.70 | 0.04 |
| **All models** |  | 0.71 | 0.71 | 0.68 | 0.03 |

**Table S4.**
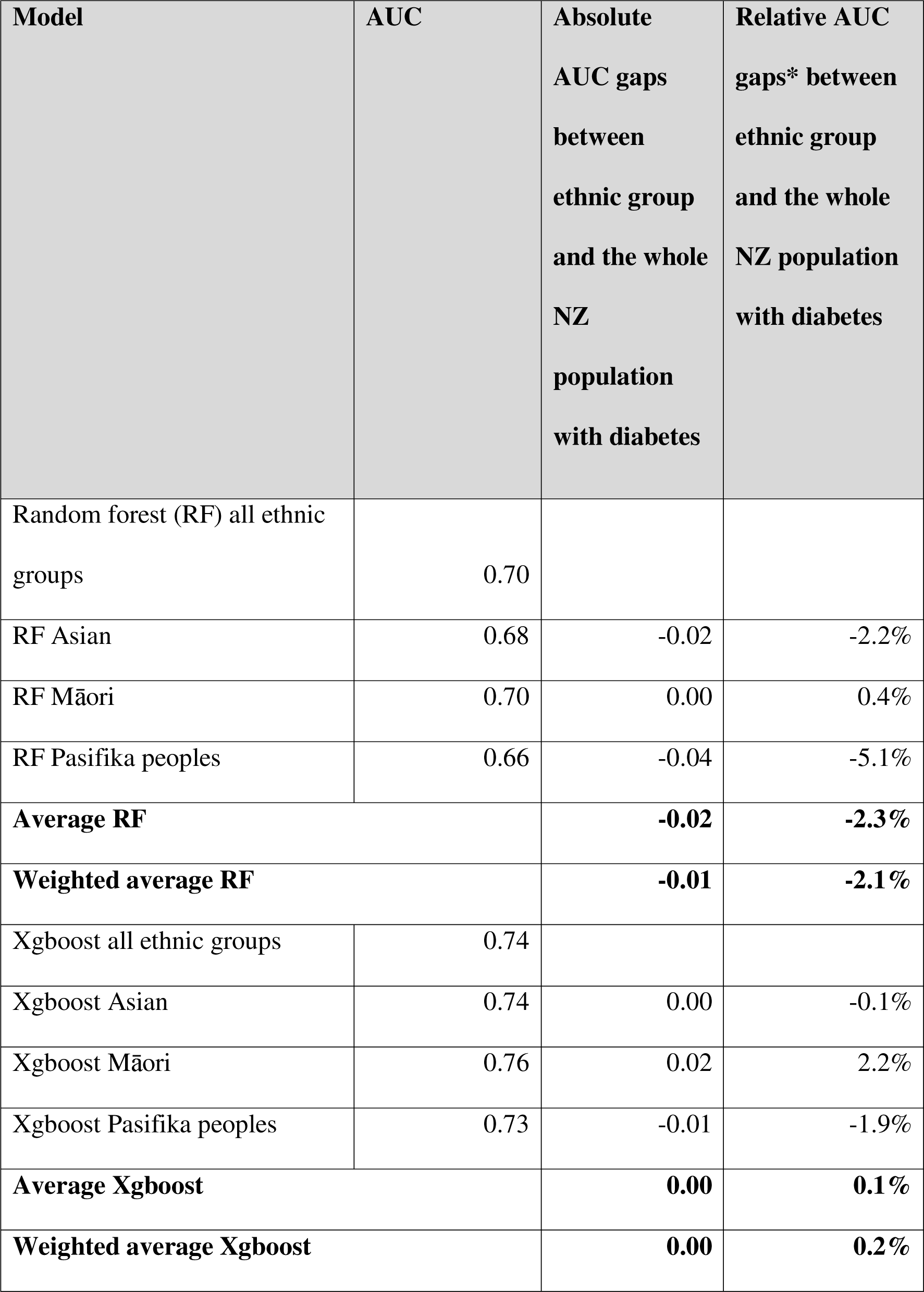
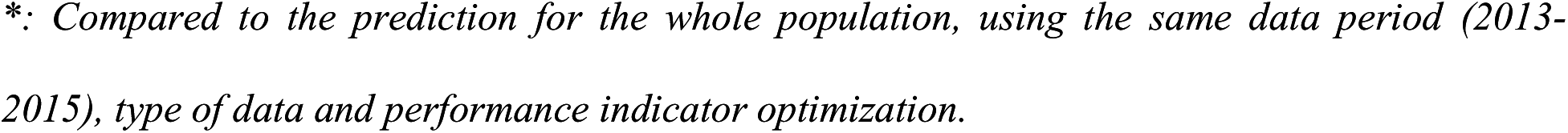
Gaps in model performance by ethnic group for the main indicator.

| **Model** | **AUC** | **Absolute**<br>**AUC gaps**<br>**between**<br>**ethnic group**<br>**and the whole**<br>**NZ**<br>**population**<br>**with diabetes** | **Relative AUC**<br>**gaps\* between**<br>**ethnic group**<br>**and the whole**<br>**NZ population**<br>**with diabetes** |
| --- | --- | --- | --- |
| Random forest (RF) all ethnic groups | 0.70 |  |  |
| RF Asian | 0.68 | -0.02 | -2.2% |
| RF Māori | 0.70 | 0.00 | 0.4% |
| RF Pasifika peoples | 0.66 | -0.04 | -5.1% |
| **Average RF** |  | **-0.02** | **-2.3%** |
| **Weighted average RF** |  | **-0.01** | **-2.1%** |
| Xgboost all ethnic groups | 0.74 |  |  |
| Xgboost Asian | 0.74 | 0.00 | -0.1% |
| Xgboost Māori | 0.76 | 0.02 | 2.2% |
| Xgboost Pasifika peoples | 0.73 | -0.01 | -1.9% |
| **Average Xgboost** |  | **0.00** | **0.1%** |
| **Weighted average Xgboost** |  | **0.00** | **0.2%** |

*\*: Compared to the prediction for the whole population, using the same data period (2013-2015), type of data and performance indicator optimization.*

